# Protective Effects of Psychiatric Medications against COVID-19 Mortality Before Vaccines

**DOI:** 10.1101/2024.09.02.24312967

**Authors:** Rodrigo Machado-Vieira, Trudy Millard Krause, Gregory Jones, Antonio L Teixeira, Lokesh Shahani, Scott Lane, Jair C. Soares, Chau N. Truong

## Abstract

**Objectives:** To investigate the relationship between neuropsychiatric medication usage and COVID-19 outcomes before COVID-19 usage.

**Methods:** This cross-sectional study used Optum’s de-identified Clinformatics® Data Mart Database to identify patients diagnosed with COVID-19 in 2020 and their psychiatric medication prescriptions in the United States. Ordered logistic regression was used to predict the likelihood of a higher COVID-19 severity level for long-term and new users. Results were adjusted for demographic characteristics and medical and psychiatric comorbidities.

**Results:** Individuals taking all psychiatric medications were less likely to have a high severity score. Overall, users of psychiatric medications were less likely to have a higher severity score than those not taking any medications. Within individual classes, the results varied across long-term and short-term users.

Conclusions

Results of the current study suggest that psychopharmacological agents are associated with reduced COVID-19 severity levels. Specifically, our results show that antidepressant medications may be associated with a protective role against COVID-19. Considering the heightened risk of severe COVID-19 outcomes associated with depression and psychoses, early treatment with antidepressants and benzodiazepines could potentially lower the incidence of severe cases and mortality rates.

## Introduction

The coronavirus pandemic in 2019 (COVID-19), which emerged in the United States in early 2020, rapidly became a national public health crisis that merits in-depth analysis of risk, prevention, and disease consequences-particularly those suffering from mental illness.

Studies have shown that psychiatric diagnoses such as schizophrenia, mood, and anxiety disorders are independent risk factors for COVID-19 infection and severity of symptoms(1,2). However, a previous study by the authors showed that while patients with schizophrenia had a higher death rate from COVID-19 than the general population, patients with schizophrenia and mood disorders had significantly lower rates of COVID-19 positivity than the general population(3). These seemingly contrasting findings raise questions as to whether various classes of psychiatric medications themselves might possess protective effects against COVID-19.

Indeed, preclinical studies have shown that certain antidepressants provide antiviral effects against SARS-CoV-2 in different models(4,5). Evidence from preliminary studies supports the potential benefit of selective serotonin reuptake inhibitors (SSRIs) in preventing more severe outcomes in COVID-19(6), which have anti-inflammatory properties by reducing the activation of pro-inflammatory cytokines such as IL-6 and TNF and by limiting the activity of the sphingomyelinase/ceramide system implicated in COVID-19 infection(7–9).

Other small pilot studies, although not all, have demonstrated that patients treated with antidepressants, such as SSRI fluvoxamine, can result in lower mortality rates and decreased COVID-19 severity (10–13). Transcriptomic analysis of the atypical antipsychotic aripiprazole also demonstrated the ability to antagonize deleterious effects induced by SARS-CoV-2 infection(14). However, other studies showed that short-term exposure to atypical antipsychotics might increase the risk of altered immune response in COVID-19(15).

The present study aimed to add clarity to this issue by investigating whether specific classes of psychiatric medications reduced the odds of infection and the severity of COVID-19 symptoms. To isolate the effects of individual medication classes more directly, we included only persons who had a history of psychotropic medication use and were subsequently diagnosed with COVID-19 between March and December of 2020, before the widespread dissemination of vaccines. Most prior studies on psychiatric medications and COVID-19 outcomes used data from a single medical record source, limiting sample size availability, power, and generalizability outside the region where the data were obtained. We analyzed administrative claims data from Optum’s de-identified Clinformatics® Data Mart Database. This dataset is derived from a database of administrative health claims for large commercial and Medicare Advantage health plan members. The population is geographically diverse, spanning all 50 states, which provides us with the advantage of studying a large and nationally representative cohort longitudinally.

Overall, we hypothesized that some classes of psychiatric medications and the duration of psychopharmacological treatment would be associated with a reduction in the relative risk of mortality and severity of illness for patients with COVID-19.

## Methods

### 1.1 Data

Data were obtained from Optum’s de-identified Clinformatics® Data Mart Database (Clinformatics®), which is derived from a database of administrative health claims for members of large commercial and Medicare Advantage health plans. Clinformatics® utilizes medical and pharmacy claims to derive patient-level enrollment information, health care costs, and resource utilization information. The population is geographically diverse, spanning all 50 states, and is statistically de-identified under the Expert Determination method, which is consistent with

HIPAA and managed according to Optum® customer data use agreements. The Clinformatics® administrative claims submitted for payment by providers and pharmacies are verified, adjudicated and de-identified prior to inclusion. The dataset was accessed through the University of Texas (UTHSC-Houston) School of Public Health Center for Health Care Data (CHCD), from January 2022 until February 2023, and the study was reviewed and approved by the UTHSC-Houston institutional review board in accordance with the Belmont Report and the Declaration of Helsinki.

COVID-19 severity levels were identified for each patient by using a modified severity scale(15) that took into account procedure and revenue codes and was adapted from the World Health Organization (WHO)’s COVID-19 Progression Index. A total of 9 levels of severity were distinguished, with level 1 assigned to unconfirmed cases which were not included in this study. For analysis, each severity level was grouped into four groups. Group 1 consisted of severity levels 2 and 3. Group 2 consisted of severity levels 4, 5, and 6. Group 3 consisted of severity levels 7 and 8, and Group 4 included only severity level 9.

### 1.2 Inclusion Criteria

Individuals 18 years or older with two years of continuous enrollment (2019–2020) were included in the analysis. The confirmation of COVID-19 was evidenced by a recorded diagnosis on a claim for healthcare services. The diagnoses were identified using the International Statistical Classification of Diseases and Related Health Problems, Tenth Revision (ICD-10), U07.1, and U07.2. The U07.2 code signifies persons for whom COVID-19 is diagnosed clinically or epidemiologically, but lab testing is inconclusive or unavailable. Only 15 eligible enrollees received this diagnostic code, all of whom eventually received a positive laboratory test to confirm COVID-19. They were thus all included in the final cohort. The earliest date an individual confirmed COVID-19 after March 1, 2020, was established as the index date. Therefore, individuals diagnosed with COVID-19 from the index date to December 31^st^, 2020, were included in the study.

Individuals who filled a prescription for the above medications were divided into three mutually exclusive groups based on their prescription information within 120 days before receiving a COVID-19 diagnosis: 1) New Users, 2) Long-Term Users, and 3) non-Users. New Users had a prescription within 30 days before having a COVID-19 diagnosis but no other prescription 31-120 days prior to COVID-19 diagnosis. Long-term Users were defined as individuals who had one or more prescriptions within 120 days before having COVID-19 but not exclusively occurring during the last 30 days prior to COVID-19 diagnosis. Finally, non-users were those who had no prescription for the listed medications within 120 days of getting a COVID-19 diagnosis. The 31-day cutoff was selected as sufficient time to demonstrate a medication effect. Multiple prescriptions for different drugs were excluded from testing study questions without the potential confound of drug-drug interactions.

### 1.3 Variables

The main variables aiming to predict outcomes were psychiatric medication type and medication duration prior to receiving a COVID-19 diagnosis. The three medication categories were antipsychotics, antidepressants, and lithium. Antidepressant medications included selective serotonin reuptake inhibitors (SSRIs), serotonin and norepinephrine reuptake inhibitors (SNRIs), phenylpiperazine, tetracyclic and tricyclic antidepressants, as well as miscellaneous antidepressants (5-hydroxytryptophan, brexanolone, bupropion, esketamine, St. John’s wort, vilazodone, and vortioxetine). Benzodiazepines were listed as an independent class. The antipsychotic medications consisted of atypical antipsychotics, phenothiazines, and miscellaneous antipsychotics (haloperidol, loxapine, molindone, and pimozide). Lithium was included as a unique category because of its unique modulatory properties (16).

Demographic covariates included age, gender, and race. Race is the name of the variable provided in the Clinformatics® dataset, including the following categories: White, Black, Hispanic, Asian, and an “other/unknown.” Likewise, gender is a variable category provided by the database and may or may not reflect self-identification. Individuals with unknown gender were excluded from the analysis. Medical covariates included mental health diagnosis (schizophrenia, mood disorders, or anxiety disorders), comorbid medical conditions (hypertension, diabetes, chronic kidney disease, ischemic heart disease, metabolic syndrome, and chronic obstructive pulmonary disease), high body mass index (BMI), and current smoker status. Individuals were flagged as having medical comorbidities if they had a diagnosis within one year before having COVID-19. ICD-10 diagnosis codes were used to identify all the comorbidities except for current smoker status (Appendix 1). The current smoker was identified using ICD-10, CPT, and Logical Observation Identifiers Names and Codes (LOINC). High BMI was defined as any BMI ≥25. While it is possible for an individual to have a comorbidity more than one year before the index date and for this comorbidity to not appear in claims data one year before their COVID-19 diagnosis, we have established one year as the look-back period.

### 1.4 Statistical Analysis

Ordered logistic regression was used to predict the likelihood of having a higher COVID-19 severity level for long-term users and new users and for long-term and new users of new medication categories. Multivariate logistic regression was then used to analyze the likelihood of being in each severity level group for long-term and new users of psychotropic medications and for long-term and new users of each medication group. The analysis for this paper was generated using SAS software, Version [9.4] of the SAS System for Windows. Copyright © 2013. SAS Institute Inc. SAS and all other SAS Institute Inc. product or service names are registered trademarks or trademarks of SAS Institute Inc., Cary, NC, USA.

## Results

Overall, there were 8,488,646 individuals in the population cohort with two full years of continuous enrollment in 2019 and 2020. Four percent (366,937) had a diagnosis of COVID-19 after March 1, 2020, and were then included in the study (Fig 1). Table 1 shows the demographic information stratified by the three medication groups. There was a significant statistical difference across all demographic elements among the three groups. Individuals on psychotropic medications (Long-term Users or New Users) were older. More females than males have been prescribed medications before being diagnosed with COVID-19. As expected, long-term users and new users had a high proportion of individuals with a mental health disorder diagnosis and medical comorbidities.

**Fig 1.**
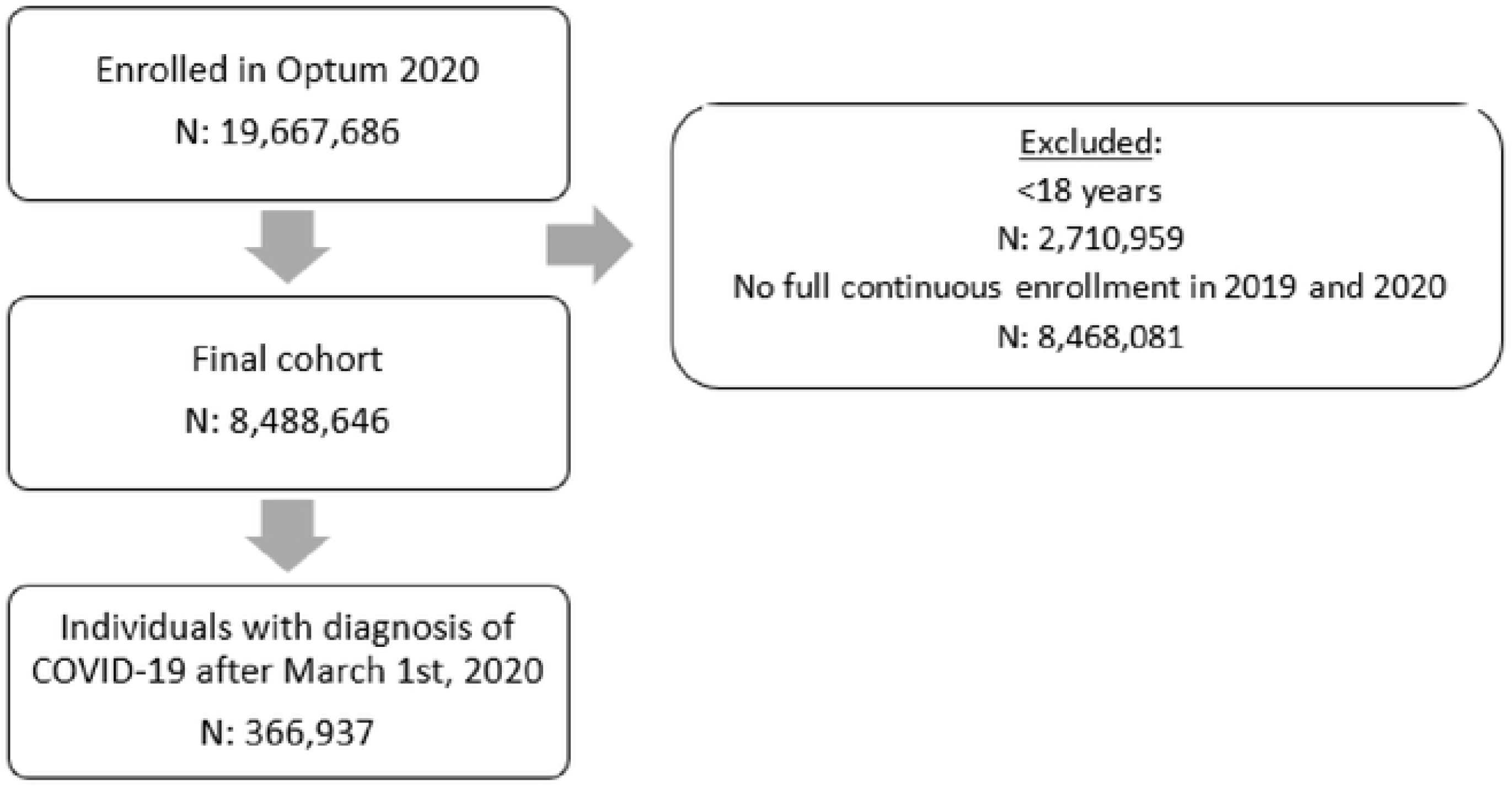
Cohort flowchart for individuals included and excluded from the present study. A total of 8,499,646 were included in the final cohort, with 366,937 having a COVID-19 diagnosis between March 1st, 2020, and December 31st, 2020

**Table 1:**
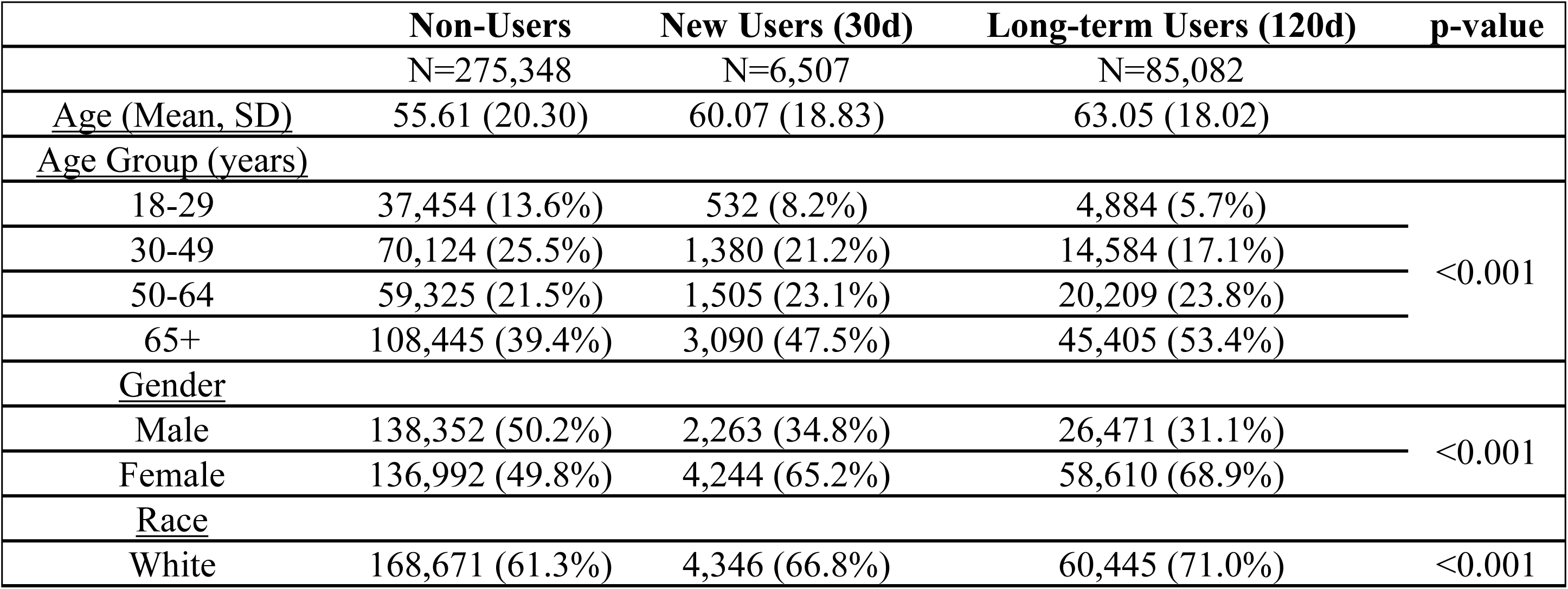
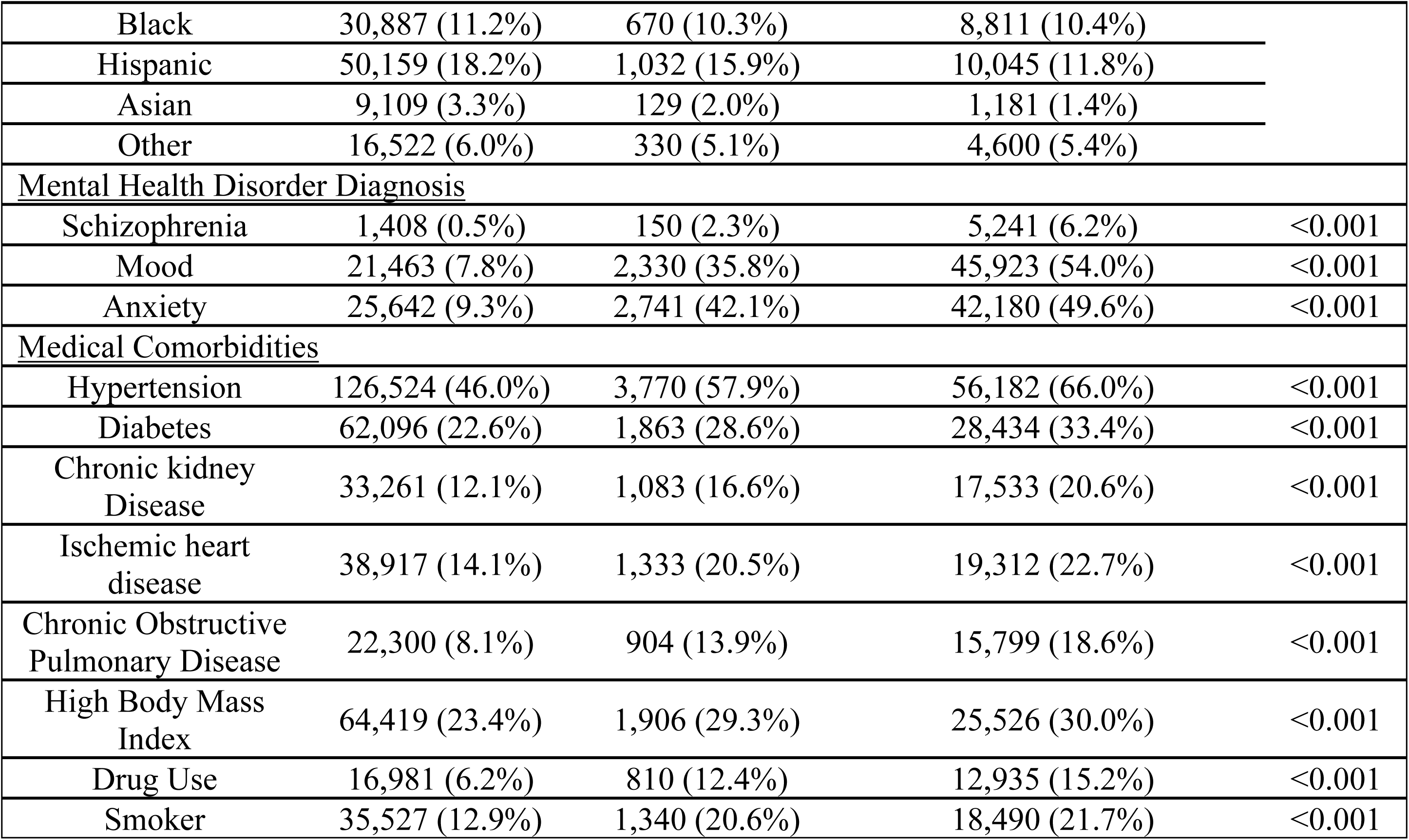
Description of Cohort.

Table 2 shows the distribution of severity levels among the three analysis groups. 75% (275,348) of individuals did not have an active medication prescription at any time within the 120 days before being diagnosed with COVID-19. However, among the 25% of individuals who did have a prescription, most had at least one prescription within 120 days before receiving a COVID-19 diagnosis. In comparison, a very small proportion of these individuals (7.1%) were new users, individuals who were newly prescribed at least one psychotropic medication within the last 30 days before having COVID-19. For all three analysis groups, most individuals resided in the lower severity levels.

**Table 2:** Distribution of Study Group among the COVID-19 Severity Levels.

| Severity | No Meds (%) | Long-Term Users (120d) (%) | New Users (30d) (%) | Total (%) |
| --- | --- | --- | --- | --- |
| 2: Ambulatory without assistance | 165,354 (60.1%) | 46,118 (54.2%) | 3,771 (58.0%) | 215,243 (58.7%) |
| 3: Ambulatory requiring assistance | 38,481 (14.0%) | 11,608 (13.6%) | 864 (13.3%) | 50,953 (13.9%) |
| 4: Emergency department visit without admission | 32,537 (11.8%) | 9,308 (10.9%) | 703 (10.8%) | 42,548 (11.6%) |
| 5: Inpatient admission with no advanced treatment or oxygen | 13,901 (5.0%) | 7,230 (8.5%) | 468 (7.2%) | 21,599 (5.9%) |
| 6: Inpatient admission with non-invasive oxygen | 8,674 (3.2%) | 3,799 (4.5%) | 255 (3.9%) | 12,728 (3.5%) |
| 7: Inpatient admission and mechanical ventilation | 12,684 (4.6%) | 5,446 (6.4%) | 333 (5.1%) | 18,463 (5.0%) |
| 8: Inpatient admission, mechanical ventilation, and renal dialysis/ECMO | 1,134 (0.4%) | 403 (0.5%) | 38 (0.6%) | 1,575 (0.4%) |
| 9: Death during hospitalization | 2,583 (0.9%) | 1,170 (1.4%) | 75 (1.2%) | 3,828 (1.0%) |
| **Total** | **275,348** | **85,082** | **6,507** | **366,937** |

After adjusting for comorbidities, mental health diagnoses, and demographic characteristics (age, race, gender, BMI), individuals taking any type of psychotropic medication were less likely to have a high COVID-19 severity score (Table 3). Long-term users were 9% less likely to have a higher severity score (CI: 0.89-0.93, p-value <0.001) than non-users. New users, those who started using psychotropic medications within 30 days before receiving a COVID-19 diagnosis, were also significantly less likely to have a higher severity score (OR: 0.90, CI: 0.86-0.96, p-value<0.001) than non-users. In the adjusted model, those who were new SSRI antidepressant users were 15% (CI: 0.79-0.93, p-value<0.001) less likely to have a higher severity score. This decrease in the risk of a higher COVID severity score can also be seen for new users of miscellaneous antidepressants and benzodiazepines. However, new users of phenothiazine antipsychotics were more likely to have a higher severity score (OR: 1.48; CI: 1.16-1.89; p-value: 0.002). Long-term users of SSRI antidepressants (OR: 0.91; CI: 0.89-0.94; p-value <0.001), miscellaneous antidepressants (OR: 0.84, CI: 0.80-0.88; p-value<0.001), and benzodiazepines (OR: 0.91; CI: 0.88-0.94; p-value<0.001) were less likely to have a higher severity score. However, taking tricyclic antidepressants and phenothiazine long-term resulted in a 19% (CI: 1.12-1.27; p-value<0.001) and 25% (CI: 1.09-1.43; p-value: 0.002) chance of having a higher COVID severity score, respectively.

**Table 3:**
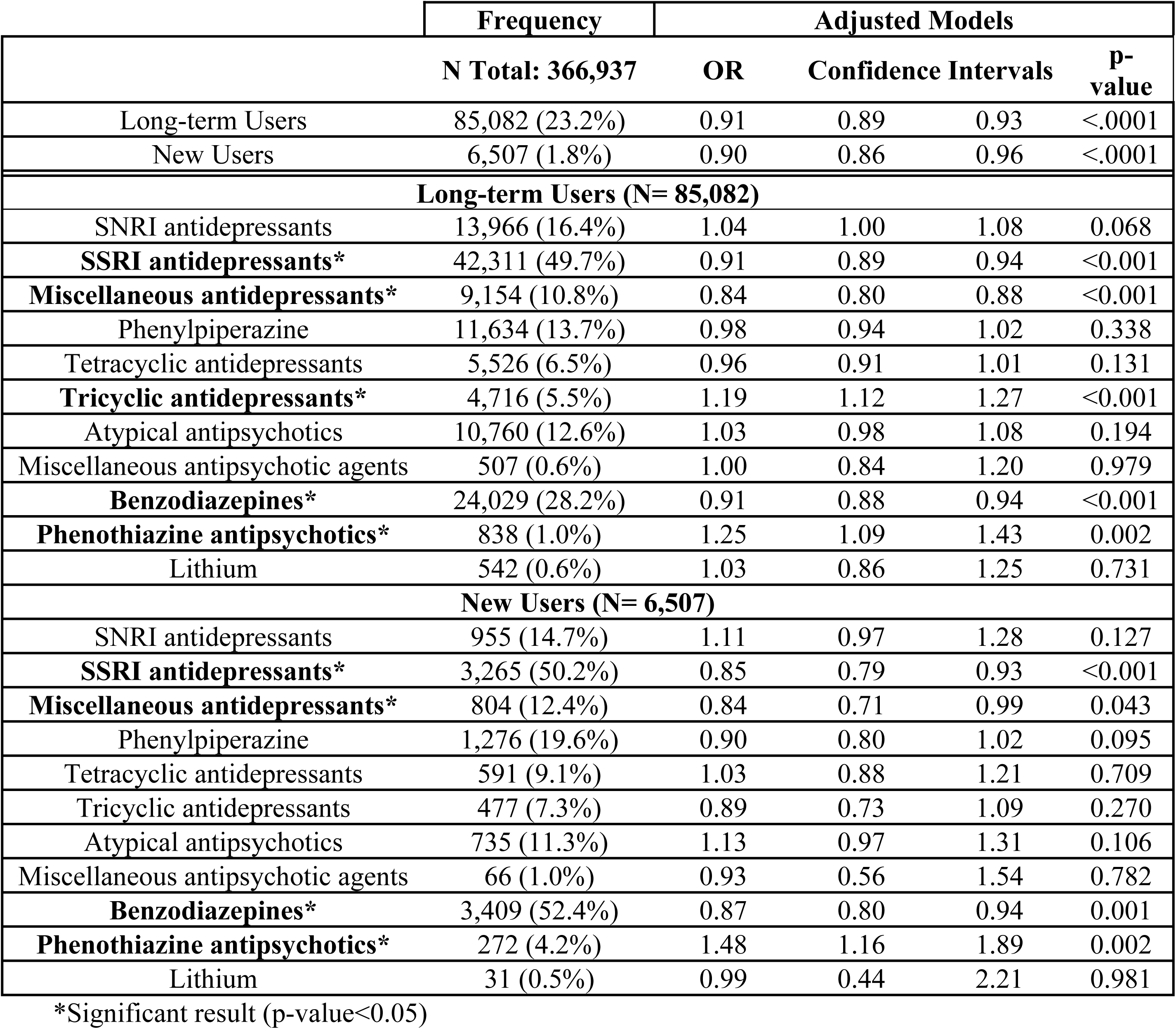
Frequency and Adjusted Odds Ratio of Having a Higher Severity Score by Medications.

Table 4 shows the adjusted odds ratio for the likelihood of having each severity group stratified by long-term users and new users. Specifically, long-term and new users of SSRI antidepressants or benzodiazepines were significantly more likely to have a severity score in Group 1 rather than a higher severity score. However, some medication classes were associated with a high risk of having a higher severity score. This can be seen among long-term tricyclic antidepressants and phenothiazine antipsychotic users and new users of phenothiazine antipsychotics. For SNRI antidepressants, new users incurred significantly higher mortality rates (OR: 1.83, CI 1.17, 2.85), and long-term users were significantly 16% more likely to be in severity group 3 (i.e., mechanical ventilation ± renal dialysis/ ECMO) (CI: 1.08,1.24). Long-term users of tricyclic antidepressants, atypical antipsychotics, and phenothiazine antipsychotics were also at a higher risk of experiencing mortality.

**Table 4:**
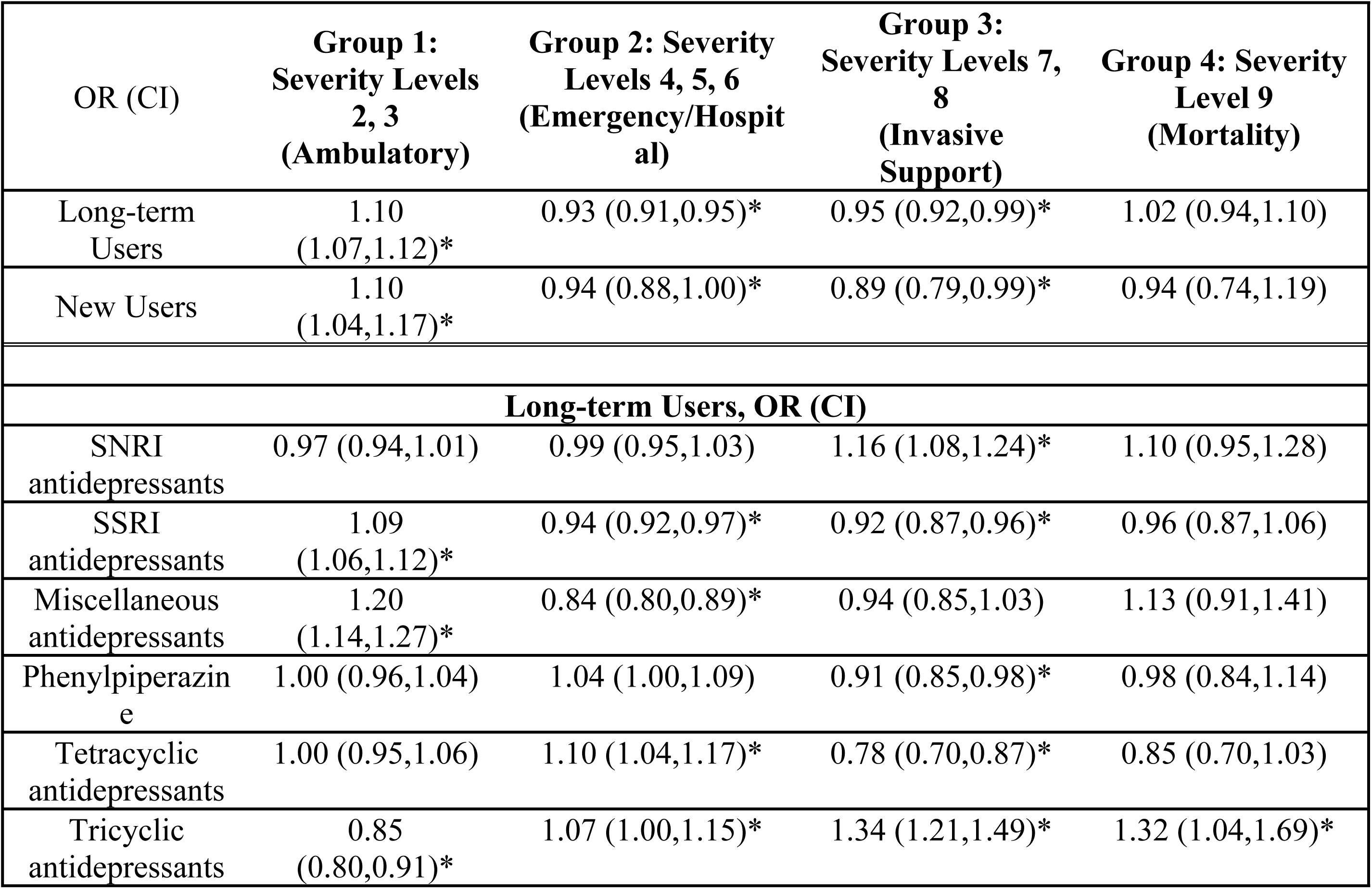
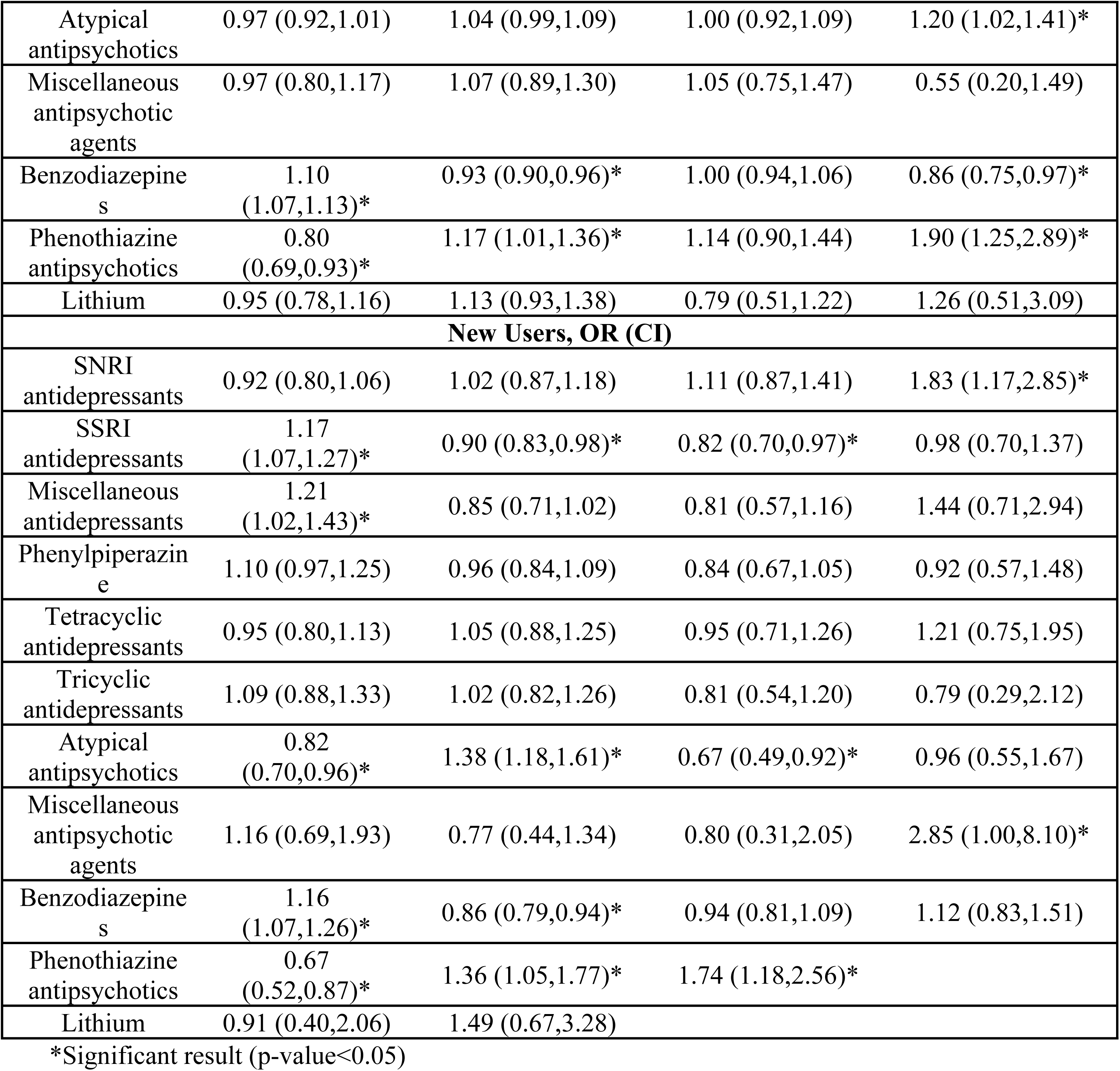
Adjusted ORs for each severity level group by medications.

| OR (CI) | Group 1:<br>Severity Levels<br>2, 3<br>(Ambulatory) | Group 2: Severity<br>Levels 4, 5, 6<br>(Emergency/Hospital) | Group 3:<br>Severity Levels 7, 8<br>(Invasive Support) | Group 4: Severity<br>Level 9<br>(Mortality) |
| --- | --- | --- | --- | --- |
| Long-term Users | 1.10<br>(1.07,1.12)* | 0.93 (0.91,0.95)* | 0.95 (0.92,0.99)* | 1.02 (0.94,1.10) |
| New Users | 1.10<br>(1.04,1.17)* | 0.94 (0.88,1.00)* | 0.89 (0.79,0.99)* | 0.94 (0.74,1.19) |
| **Long-term Users, OR (CI)** |  |  |  |  |
| SNRI antidepressants | 0.97 (0.94,1.01) | 0.99 (0.95,1.03) | 1.16 (1.08,1.24)* | 1.10 (0.95,1.28) |
| SSRI antidepressants | 1.09<br>(1.06,1.12)* | 0.94 (0.92,0.97)* | 0.92 (0.87,0.96)* | 0.96 (0.87,1.06) |
| Miscellaneous antidepressants | 1.20<br>(1.14,1.27)* | 0.84 (0.80,0.89)* | 0.94 (0.85,1.03) | 1.13 (0.91,1.41) |
| Phenylpiperazine | 1.00 (0.96,1.04) | 1.04 (1.00,1.09) | 0.91 (0.85,0.98)* | 0.98 (0.84,1.14) |
| Tetracyclic antidepressants | 1.00 (0.95,1.06) | 1.10 (1.04,1.17)* | 0.78 (0.70,0.87)* | 0.85 (0.70,1.03) |
| Tricyclic antidepressants | 0.85<br>(0.80,0.91)* | 1.07 (1.00,1.15)* | 1.34 (1.21,1.49)* | 1.32 (1.04,1.69)* |
| Atypical antipsychotics | 0.97 (0.92,1.01) | 1.04 (0.99,1.09) | 1.00 (0.92,1.09) | 1.20 (1.02,1.41)* |
| Miscellaneous antipsychotic agents | 0.97 (0.80,1.17) | 1.07 (0.89,1.30) | 1.05 (0.75,1.47) | 0.55 (0.20,1.49) |
| Benzodiazepines | 1.10 (1.07,1.13)* | 0.93 (0.90,0.96)* | 1.00 (0.94,1.06) | 0.86 (0.75,0.97)* |
| Phenothiazine antipsychotics | 0.80 (0.69,0.93)* | 1.17 (1.01,1.36)* | 1.14 (0.90,1.44) | 1.90 (1.25,2.89)* |
| Lithium | 0.95 (0.78,1.16) | 1.13 (0.93,1.38) | 0.79 (0.51,1.22) | 1.26 (0.51,3.09) |
| **New Users, OR (CI)** |  |  |  |  |
| SNRI antidepressants | 0.92 (0.80,1.06) | 1.02 (0.87,1.18) | 1.11 (0.87,1.41) | 1.83 (1.17,2.85)* |
| SSRI antidepressants | 1.17 (1.07,1.27)* | 0.90 (0.83,0.98)* | 0.82 (0.70,0.97)* | 0.98 (0.70,1.37) |
| Miscellaneous antidepressants | 1.21 (1.02,1.43)* | 0.85 (0.71,1.02) | 0.81 (0.57,1.16) | 1.44 (0.71,2.94) |
| Phenylpiperazine | 1.10 (0.97,1.25) | 0.96 (0.84,1.09) | 0.84 (0.67,1.05) | 0.92 (0.57,1.48) |
| Tetracyclic antidepressants | 0.95 (0.80,1.13) | 1.05 (0.88,1.25) | 0.95 (0.71,1.26) | 1.21 (0.75,1.95) |
| Tricyclic antidepressants | 1.09 (0.88,1.33) | 1.02 (0.82,1.26) | 0.81 (0.54,1.20) | 0.79 (0.29,2.12) |
| Atypical antipsychotics | 0.82 (0.70,0.96)* | 1.38 (1.18,1.61)* | 0.67 (0.49,0.92)* | 0.96 (0.55,1.67) |
| Miscellaneous antipsychotic agents | 1.16 (0.69,1.93) | 0.77 (0.44,1.34) | 0.80 (0.31,2.05) | 2.85 (1.00,8.10)* |
| Benzodiazepines | 1.16 (1.07,1.26)* | 0.86 (0.79,0.94)* | 0.94 (0.81,1.09) | 1.12 (0.83,1.51) |
| Phenothiazine antipsychotics | 0.67 (0.52,0.87)* | 1.36 (1.05,1.77)* | 1.74 (1.18,2.56)* |  |
| Lithium | 0.91 (0.40,2.06) | 1.49 (0.67,3.28) |  |  |

\*Significant result (p-value<0.05)

## Discussion

The main findings of this study were that subjects taking different classes of psychiatric medications showed a lower risk for severe COVID-19 symptomatology after adjusting for demographic variables, psychiatric diagnosis, and other medical comorbidities. Studies have suggested that psychiatric diagnosis may be an independent risk factor for COVID-19 infection(2,17). After adjustment, a lower likelihood of having a higher severity score was seen in both short and long-term users of SSRIs, miscellaneous antidepressants, and benzodiazepines. The use of phenothiazine antipsychotics for any duration appeared to confer more severe illness, as well as long-term use of tricyclics. Such findings may be of relevance for future pandemics, especially with regard to acute stabilization.

Regarding classification strategies for severity, many confounding factors exist in stratifying patients at individual severity levels. For example, local hospital bed availability, individual emergency department bandwidth and criteria for discharge, patient capacity/resources to seek ambulatory care before progression of illness, and other factors may influence where patients end up (i.e., level 3 versus 5). We thus stratified the nine coded levels into four groupings (ambulatory, emergency/inpatient, invasive support (i.e., mechanical ventilation/ECMO/dialysis), and in-hospital mortality). We suggest that this approach can control for some of the non-illness-related influences mentioned above and present a more logical representation of the data.

Our results regarding SSRI and various other antidepressant use coincide with previous literature, which suggested that taking these agents may reduce the severity of COVID-19’s direct effects on the immune system, in part by suppression of pro-inflammatory cytokine signaling (18,19), modulation of clathrin-mediated viral endocytosis(20). and other still mechanisms. Overall, evidence suggests that SSRIs, especially fluoxetine, can inhibit the replication of a wide range of viruses *in vitro*, including SARS-CoV-2(4,21,22). It was also shown that there is a 27–57% risk decrease (HR, 0.56; 95% CI, 0.43–0.73, *p* < 0.001) of intubation or COVID-19-related death in subjects treated with antidepressants(12,23). The SSRI Fluvoxamine induced improvement of clinical outcomes in symptomatic COVID-19(10,24). Also, the use of antidepressants was associated with a reduced risk of intubation or death in hospitalized patients with COVID-19(21,22).

One prominent candidate pathway may involve inhibiting the acid sphingomyelinase/ceramide system. Upregulation of this system by viral engagement is thought to increase the formation of ceramide-enriched membrane domains facilitating viral entry via clustering of angiotensin-converting enzyme 2 (ACE-2) binding domains for viral particles (7,21,22,25). Indeed, plasma ceramides have been shown to correlate strongly with clinical severity and markers of inflammation in patients with COVID-19(26). As follows, many antidepressants (i.e., most tricyclic antidepressants, duloxetine, escitalopram, fluoxetine, fluvoxamine, sertraline, and others) and certain antipsychotics (i.e., aripiprazole, phenothiazines, and others) may be considered functional inhibitors of acid sphingomyelinase (FIASMA), with potential utility in COVID-19(22,24). Contrasting findings in our study were demonstrated with regard to FIASMA medications. That is, protective associations with SSRIs but increased severity and/or mortality associated with SNRIs, tricyclics, and phenothiazines. A possible explanation for this is the opposing adverse effects seen with many of these medications. For example, anticholinergic properties demonstrated by many of these medications have been repeatedly associated with increased risk and/or severity of pneumonia(27). Likewise, cardiotoxic effects (QTc prolongation, sodium channel blockade) with phenothiazines and tricyclics in particular(28) may override the influence of sphingomyelinase inhibition, increasing the risk of cardiac decompensation in patients already hospitalized with COVID-19.

Regarding mortality risk (group 4/level 9) specifically, our subgroup analysis found an increased likelihood of death with long-term use of tricyclic antidepressants, atypical antipsychotics, and phenothiazine antipsychotics. A reduction in the likelihood of mortality was seen with long-term use of benzodiazepines. Assessment of mortality risk in our sample should be tempered in the context of relatively small sample sizes (approximately 1% of each group and total study). Indeed, mixed findings have been demonstrated regarding benzodiazepine use and COVID-19 mortality (29,30). Hoertel et al. demonstrated an increased risk for most agents but a decreased mortality risk with diazepam(30). Park (2022) has demonstrated that the use of benzodiazepines among South Korean COVID-19 patients was associated with an increased risk of hospitalization but not COVID-19 seropositivity, severe outcomes, or mortality (28). Baseline differences in the population studied, as well as differential classification of short-term (90 days) and chronic use (180 days), may have contributed to some of these differences as well.

Overall, findings within the various antipsychotic classes highlight the complex, multifactorial nature of COVID-19 severity in relation to medication effects. As multi-medication users were excluded from our study, this cohort excludes those with unipolar depression using atypical medication for augmentation. Thus, behavioral and biological factors related to psychotic or bipolar illness may also contribute to severity and mortality risk, in addition to any medication effects(31). For example, newly initiated atypical antipsychotics were associated with an increased risk of hospitalization or emergency department visits (group 2) but reduced risk of invasive treatment (mechanical ventilation, dialysis, and ECMO (group 3) and no change in mortality. This may suggest a short-term protective effect of this class, which shares overlapping properties with many of the antidepressants found to be protective against COVID-19 (i.e., serotonergic and alpha-adrenergic modulation). Conversely, increased mortality risk seen with long-term usage of atypical antipsychotics may be reflective of the increased metabolic burden caused by many of these agents. Furthermore, new users of miscellaneous antipsychotics (haloperidol, loxapine, molindone, and pimozide) also demonstrated increased mortality risk— which was not seen for long-term users of these agents. Given the wide range of affordable, more tolerable alternatives in this insured population, the use of these agents may suggest severe onset of psychosis, significant decompensation, and/or highly-refractory psychotic illness—which may also increase the risk of mortality.

Limitations include the cross-sectional, retrospective design and use of data submitted by commercial plans within the United States. The findings of this study may not necessarily translate to other countries or individuals who are uninsured or covered by programs such as Medicaid. Indeed, approximately 20% of people with mental disorders are uninsured compared to 15% of the US population(32). Additionally, if death occurred outside the hospital, it was not documented in the health claims records. There is no way to assess whether rates of missing outside mortality data would differ between psychiatric medication users and non-users. The study was also limited to a select group of mood stabilizers and did not include valproate and other anti-epileptic drugs, which are also used for treating mood disorders.

Though we have controlled for proxies such as comorbid illness, BMI, and smoking, other behavioral covariates (i.e., medication compliance, quantity of healthcare interactions, alcohol consumption, social isolation, diet, exercise, etc.) not captured here may also mediate the relationship between psychiatric medication usage and COVID-19 severity/mortality (33).

Medication non-adherence is a limitation to all population-based studies. Despite variability between studies, data generally suggest comparable rates of non-adherence in patients with psychiatric disorders compared to other long-term medical conditions (i.e., 50% or more)(34,35). Non-adherence may arguably result in underestimating the magnitude of both protective effects and risk. However, the confounding influence of nonadherence to medications for other chronic medical illnesses also limits the ability to draw conclusions regarding the magnitude of its influence with certainty. Strengths include the largest known COVID-19 dataset stratified based on diverse patient groups and the adjustment for important confounders in the analyses, such as psychiatric diagnosis, medical comorbidities, and demographic characteristics. Data was also collected before the widespread dissemination of vaccines. This decision was made based on the ability to isolate any medication-related effects more directly. However, subsequent changes in population immunity and circulating variants may limit the generalizability of these findings.

## Data Availability

The data cannot be shared publicly as it belongs to a third party, Optum. The Optum® Clinformatics® Data Mart (CDM) provides richly detailed longitudinal information through statistically de-identified patient data sourced from multiple sources. However, at the time of this submission, the researchers no longer have an active contract with Optum to access the data. Those interested in using the data must contact Optum directly to gain access.

## Supporting Information

S1: Codes used to identify mental health disorder diagnosis and medical comorbidities

